# Environmental exposures associated with behavioral outcomes and their link to endogenous metabolites in children born very preterm

**DOI:** 10.64898/2026.09.25.26364017

**Authors:** Priyadarshni Patel, Youran Tan, Donghai Liang, Marie Camerota, Carmen Marsit, Barry Lester, Todd Everson

## Abstract

**Background:** Children born very preterm (VPT) are at elevated risk for adverse neurobehavioral outcomes and are vulnerable to environmental exposures. We used untargeted metabolomics analysis to identify environmentally relevant metabolites associated with childhood behavioral problems and to characterize related endogenous metabolic pathways.

**Methods:** We studied 226 children born <30 weeks gestation in a multi-site cohort study. Urine collected at age 4 was profiled using untargeted ultra-high-performance liquid chromatography high-resolution mass spectrometry (UHPLC-HRMS). Associations between 30 exogenous environmental metabolites and behavioral problems score at age 4 were evaluated cross-sectionally using multivariable linear regression, followed by metabolome-wide association and pathway enrichment analyses.

**Results:** Three exogenous metabolites were nominally associated with child behavior problems. Specifically, 2-(Acetylamino)-3-(2henylthiol)propanoic acid was positively associated with overall (β=1.42, p=0.006), externalizing (β=1.50, p=0.006), and internalizing (β=1.34, p=0.021) behavior problems. Diethofencarb was positively associated with overall (β=0.34, p=0.018) and externalizing (β=0.39, p=0.010) behavior problems, while protocatechuic acid was inversely associated with internalizing problems (β=−0.68, p=0.027). No associations remained significant after false discovery rate (FDR) correction. Nevertheless, these metabolites were associated with 151, 131, and 530 endogenous metabolic features, respectively, at FDR<0.05, highlighting perturbations in lipid metabolism, mitochondrial energetics, inflammatory signaling, amino acid metabolism, and neuroactive pathways.

**Conclusions:** Environmental metabolites associated with behavior problems were linked to broad endogenous metabolic differences relevant to neurodevelopment. These findings highlight potentially modifiable environmental influences, including exposures related to combustion-derived pollutants, pesticides, and dietary sources, that may contribute to behavioral vulnerability among children born very preterm.

## Introduction

Early childhood is a critical window for brain development, during which environmental exposures can have lasting effects on neurobehavioral outcomes [1]. The developing brain is particularly vulnerable to chemical exposures from the environment, including pollutants, dietary compounds, and lifestyle-related chemicals, which may influence biological pathways such as oxidative stress, inflammation, and endocrine disruption [2]. These mechanisms may be especially relevant among children born very preterm (<30 weeks of gestation), who are at increased risk for neurodevelopmental and behavioral difficulties, including higher levels of internalizing and externalizing behavior problems during childhood [3]. Understanding environmental factors that contribute to variability in these behavioral outcomes is therefore particularly important in this vulnerable population. Children born very preterm also experience a unique neonatal environment, including repeated painful procedures, mechanical ventilation, oxygen therapy, medication exposures, disrupted sleep-wake cycles, and altered sensory stimulation, which can contribute to inflammation, oxidative stress, and dysregulation of neurodevelopmental processes [4]. These features of very preterm birth may contribute to biological and neurobehavioral vulnerability, underscoring the importance of examining the adverse effects of environmental neurotoxicants in this high-risk population.

Prenatal and/or childhood exposures to common environmental toxicants, including endocrine-disrupting chemicals[5], heavy metals [6], pesticides [7], and air pollutants [8], have been associated with children’s neurodevelopmental impairments and behavioral problems. Additionally, research has linked exposure to other environmental chemicals, including phthalates, phenols, and pesticides, with adverse neurodevelopmental and behavioral outcomes in children [9, 10]. A recent review found that the majority of studies evaluating prenatal and early childhood exposures to heavy metals, endocrine-disrupting chemicals, and pesticides reported associations with adverse mental health and behavioral outcomes, including increased internalizing and externalizing behaviors [2]. Together, these findings suggest that behavioral development is likely influenced by a broad range of environmental chemicals, many of which remain incompletely characterized for neurodevelopmental effects. This knowledge gap may be particularly important among children born very preterm, for whom the environmental contaminants encountered during childhood and their potential neurobehavioral effects remain understudied. Given the heightened biological vulnerability associated with very preterm birth, identifying environmental exposures that may contribute to behavioral development in this population is especially important.

Additionally, evidence from experimental and epidemiological studies suggests that environmental chemical exposures may influence neurodevelopment not only through direct neurotoxic effects but also by inducing broad disruptions in metabolic and physiological processes that are essential for brain development [11]. For example, EDCs can perturb mitochondrial function, energy metabolism, lipid homeostasis, and redox balance, leading to systemic metabolic dysregulation and chronic low-grade inflammation [12]. Disruptions in mitochondrial and cellular energy metabolism have been implicated in neurodevelopmental disorders and are thought to contribute to altered neuronal signaling, impaired synaptic plasticity, and long-term behavioral dysfunction [13]. Biological disruptions may be particularly consequential for children born very preterm, who experience heightened vulnerability in biological systems involved in neurodevelopment, including inflammatory, oxidative stress, and metabolic processes. Despite this vulnerability, little is known about the biological pathways through which environmental exposures may influence behavioral development in children born very preterm.

Metabolomics offers a powerful approach to simultaneously capture both endogenous metabolites that reflect underlying physiological and metabolic processes, as well as exogenous chemical and their metabolic products that can provide insights into environmental exposures. Metabolome-wide association studies (MWAS) are data driven approaches to systematically evaluate associations between metabolites and health outcomes. MWAS allows for the analysis of hundreds to thousands of metabolites at once, enabling the identification of exposure-related biomarkers and their links to phenotypic outcomes. This approach has been increasingly applied in environmental health research to identify metabolic signatures of chemical exposures and to identify potential biological pathways involved in disease processes [14, 15]. Applying metabolomics to the study of VPT children provides an opportunity to broadly characterize environmental exposures during childhood while simultaneously identifying the biological perturbations that may link these exposures with behavioral development.

The current study leverages data from the Neonatal Neurobehavior and Outcomes in Very Preterm Infants (NOVI) Study, a multicenter cohort of children born very preterm. Using an untargeted metabolomics framework, we aimed to identify environmentally relevant metabolites associated with behavioral problems at age 4 and to characterize endogenous metabolic pathways linked to these exposures. We first evaluated the associations between environmentally relevant metabolites and parent-reported total, internalizing, and externalizing behavioral problems, as measured by Behavior Assessment System for Children (BASC-3). We then conducted MWAS to identify the endogenous metabolites and metabolic pathways potentially linked to these behavior-associated environmentally-relevant metabolites.

## Methodology

### Study Population

Participants were drawn from the Neonatal Neurobehavior and Outcomes in Very Preterm Infants (NOVI) Study, a multi-site study of infants born < 30 weeks gestational age (GA). Participants were recruited from nine university affiliated NICUs across six research sites from April 2014 to June 2016. Inclusion criteria were: (1) birth < 30 weeks GA, (2) parental ability to speak English or Spanish, (3) residence within 3 hours of the NICU and follow-up clinic. Exclusion criteria included major congenital anomalies, maternal age < 18 years, maternal cognitive impairment, and infant death. Parents of eligible infants were approached when infants were 31-32 weeks GA or when survival to discharge was deemed likely by the attending neonatologist. Each site obtained informed consent in line with its institution’s review board. Of the 704 infants enrolled at birth, 393 participants attended a follow-up visit at age 4. Of these, 226 provided urine samples for metabolomics profiling and completed the behavior problem assessment and were included in this study.

### Behavioral Assessment

Behavioral outcomes were assessed at age 4 using the Behavior Assessment System for Children (BASC), which captures multiple domains including externalizing and internalizing behaviors [16, 17]. The BASC [18] is a caregiver-report instrument designed to assess adaptive and maladaptive behaviors in children aged 2 to 5 years across home and community contexts. Parents rate 139 behaviorally descriptive items using a 4-point Likert scale ranging from 0 (“Never”), 1 (“Sometimes”), 2 (“Often”), to 3 (“Almost Always”). Items are organized into 12 primary subscales: Hyperactivity, Aggression, Anxiety, Depression, Somatization, Attention Problems, Atypicality, Withdrawal, Adaptability, Social Skills, Activities of Daily Living, and Functional Communication. These subscales are further aggregated into four composite domains. Externalizing Problems is derived from Hyperactivity, Aggression, and Conduct Problems subscales; Internalizing Problems comprises Anxiety, Depression, and Somatization subscales; the Behavioral Symptoms Index (BSI) integrates Externalizing and Internalizing composites along with Attention Problems, Atypicality, and Withdrawal; and Adaptive Skills combines Adaptability, Social Skills, Activities of Daily Living, and Functional Communication. Raw scores for each subscale and composite are summed and converted into standardized T-scores for interpretation. T-scores between 60-69 are classified as “at risk,” while scores of 70 or above indicate “clinically significant” concerns. The BASC-3 Parent Rating Scale–Preschool (PRS-P) has demonstrated strong internal consistency and test-retest reliability [19].

### Ultra-performance liquid chromatography along with high-resolution mass spectrometry (UPLC-HRMS)

Untargeted metabolomic profiling was conducted by the North Carolina Human Health Exposure Analysis Resource (HHEAR) Hub at the Sumner Metabolomics and Exposome Laboratory (University of North Carolina at Chapel Hill Nutrition Research Institute). Biospecimens were analyzed using ultra-high performance liquid chromatography coupled to high-resolution mass spectrometry (UHPLC-HRMS) on a Thermo Fisher Q Exactive HF-X mass spectrometer (Serial No. SN06588L), following previously published methods[20] [21]. Data were acquired in positive ionization mode.

Raw LC-MS data were processed for feature detection and alignment, and compound annotation was performed using the ADAP-KDB workflow. Each detected signal was assigned a unique signal identifier, retention time, precursor m/z, adduct, and neutral mass where applicable. When multiple adducts were present, retention time was averaged across adducts and precursor ions were ordered by intensity. Putative compound identification was based on matching against spectral libraries using accurate mass, retention time error, isotopic similarity, and MS/MS fragmentation similarity (both experimental and theoretical). Annotation confidence was assigned using ontology-based matching criteria as implemented in the ADAP-KDB framework [22].

To ensure data quality, background signals were removed by excluding features in which the mean intensity in blank samples exceeded the mean intensity in study pooled quality control samples, applied batch-by-batch. Blanks consisted of LC-MS grade water, and pooled QC samples were generated by the NC HHEAR laboratory. Additional filtering was applied by excluding features with a relative standard deviation (RSD) greater than 50% across QC samples.

Data normalization was performed using total ion intensity normalization followed by batch effect correction using previously established methods [23] and data was log10 transformed. The resulting dataset was used for downstream statistical analyses. Compound annotation outputs included retention time, precursor m/z, adduct type, neutral mass, retention time error, precursor mass error (ppm), isotopic similarity, MS/MS fragmentation scores (experimental and theoretical), and ontology-level confidence scores [22].

### Chemical identification and annotation

Metabolomic signals were identified by comparison with the NC HHEAR Hub’s IESL, which includes more than 2,400 compounds comprising both endogenous and exogenous metabolites. Endogenous metabolites reflect processes related to an individual’s metabolism, whereas exogenous metabolites represent lifetime exposures, including environmental contaminants, dietary compounds, pharmaceuticals, and secondary metabolites generated following exposure. All standard reference compounds included in the IESL were analyzed under the same conditions as the study samples. Metabolite identification and annotation were evaluated using ontology levels (OLs), which indicate the strength of evidence supporting each assignment. These levels were determined based on retention time (RT), exact mass (MS), MS/MS fragmentation patterns, and isotopic ion patterns. Signals that closely matched compounds in the IESL by RT (within ±0.5 min), MS (<5 ppm), and MS/MS fragmentation patterns (similarity score >30) were assigned an OL1 designation. Signals matching the IESL by RT and MS were classified as OL2a. Metabolites assigned OL1 or OL2a were considered confidently identified in this study. Signals that matched the IESL by MS and MS/MS fragmentation but fell outside the specified RT window (±0.5 min) were classified as OL2b. These signals often represented conjugates or isomers with structural similarities to the corresponding compounds in the IESL.

### Statistical Analysis

All statistical analyses were performed using regression modeling frameworks. For analyses of environmental metabolites and behavioral outcomes, generalized linear regression models were fitted to evaluate associations between the feature intensity of each individual environmental metabolite (independent variables; n = 30) and BASC-3 T-scores (overall, externalizing, and internalizing domains as dependent variables). Models were adjusted based on prior literature for potential confounders including race, sex, socioeconomic status, study site, gestational age at birth and urinary creatinine to account for confounding by urinary dilution. Multiple testing correction was applied to each analysis using the false discovery rate (FDR), and adjusted p-values (q-values) < 0.05 were considered statistically significant.

To investigate downstream association with endogenous metabolites, MWAS were conducted using linear models to assess associations between the behavior-associated environmental metabolites (independent variables) and endogenous metabolite features (dependent variables; n = 16,958). All MWAS models were adjusted for the same covariates as above with the addition of 10 principle component analysis. Multiple testing correction was applied using FDR, and adjusted p-values (q-values) < 0.05 were considered statistically significant.

Pathway enrichment analyses were performed using the Metapone package [24], which uses metabolite set enrichment methods for untargeted metabolomics data. Briefly, significantly associated endogenous metabolites (FDR<0.05) were mapped to metabolic pathways, and enrichment was tested. Separate pathway enrichment analyses were performed for each set of exposure-focused MWAS results. Statistical significance was assessed using FDR correction, with pathways considered significantly enriched at adjusted p-values < 0.05. All the analyses were performed in R (version 4.5.2).

## Results

### Demographics

The study included 226 NOVI participants (Table 1), balanced by sex, with 114 females (50.4%) and 112 males (49.6%). Participants were racially diverse, with 46.5% identifying as White, 15.0% as Black, 4.9% as Asian, 0.9% as Native Hawaiian or Other Pacific Islander, and 23.9% reporting more than one race; race was unknown for 8.8% of participants. Approximately 8% of the sample were considered low SES, based on Hollingshead lowest SES category. The mean gestational age at birth was 27.24 ± 1.82 weeks.

**Table 1.**
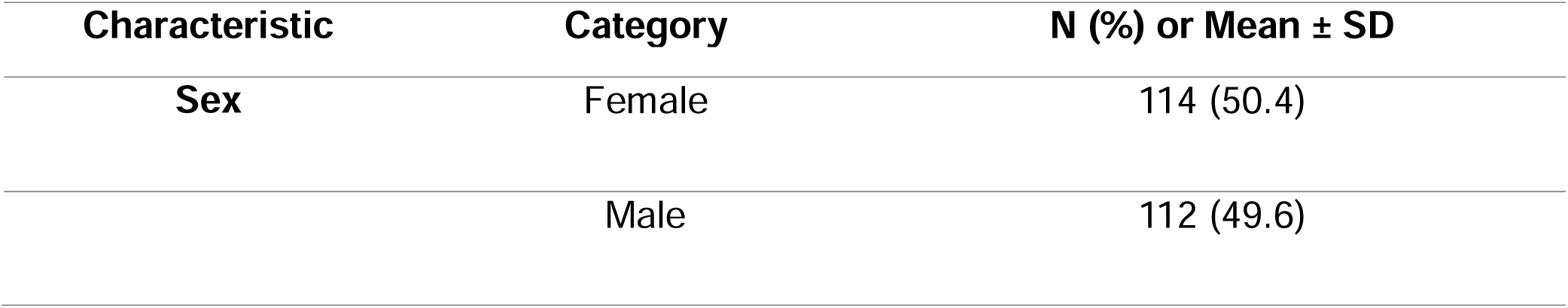
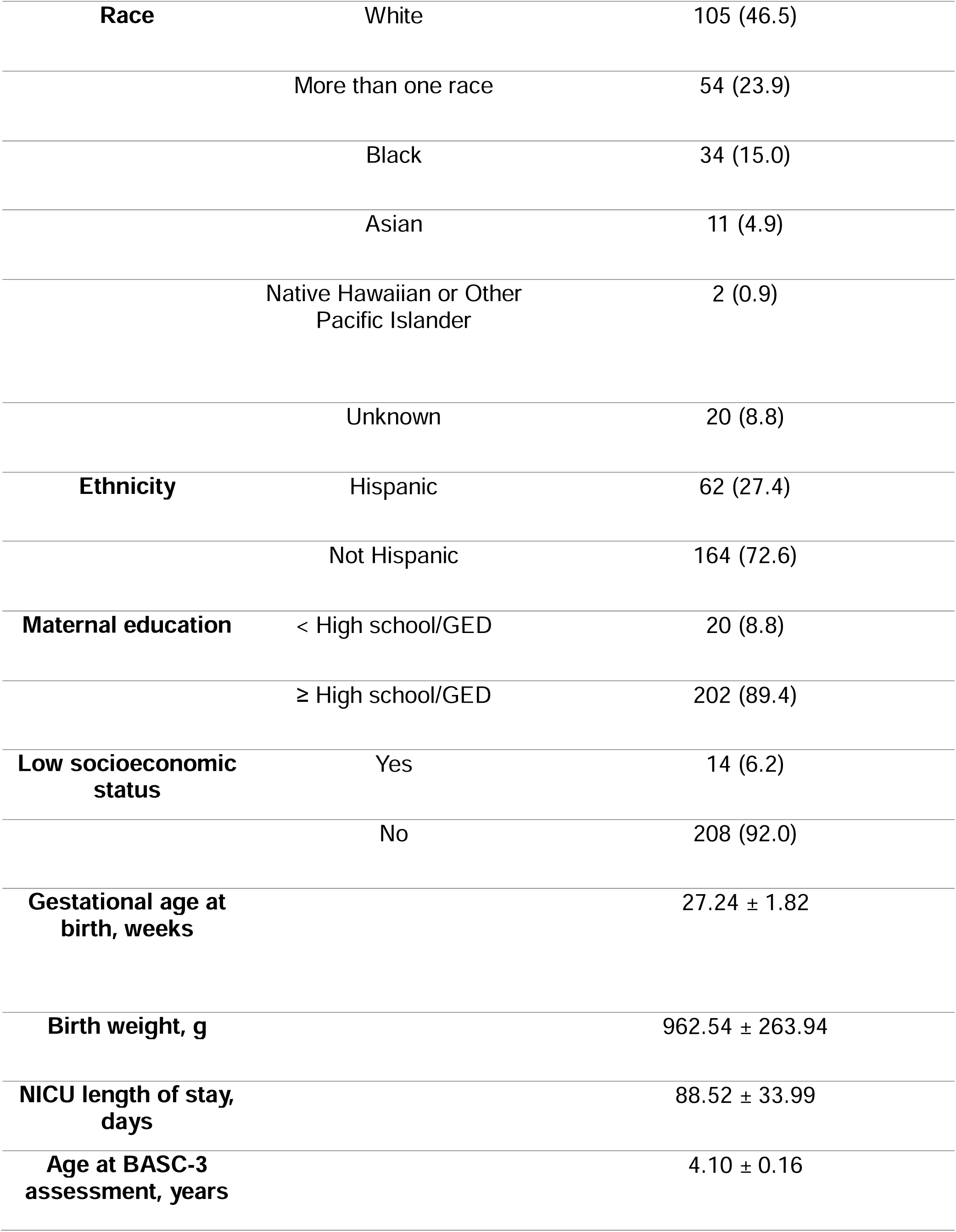
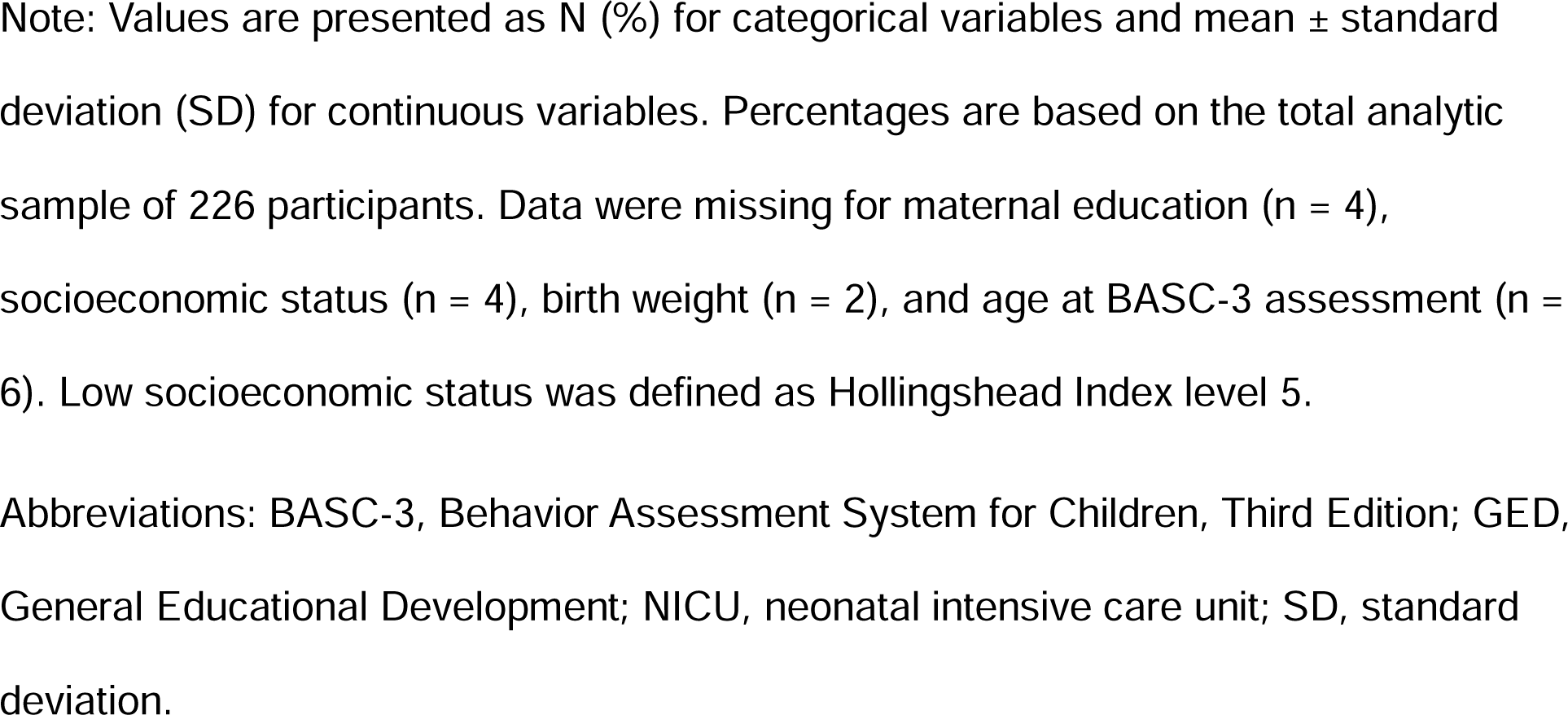
Demographic and clinical characteristics of study participants (N = 226)

| Characteristic | Category | N (%) or Mean ± SD |
| --- | --- | --- |
| Sex | Female | 114 (50.4) |
|  | Male | 112 (49.6) |
| **Race** | White | 105 (46.5) |
|  | More than one race | 54 (23.9) |
|  | Black | 34 (15.0) |
|  | Asian | 11 (4.9) |
|  | Native Hawaiian or Other<br>Pacific Islander | 2 (0.9) |
|  | Unknown | 20 (8.8) |
| **Ethnicity** | Hispanic | 62 (27.4) |
|  | Not Hispanic | 164 (72.6) |
| **Maternal education** | < High school/GED | 20 (8.8) |
|  | ≥ High school/GED | 202 (89.4) |
| **Low socioeconomic**<br>**status** | Yes | 14 (6.2) |
|  | No | 208 (92.0) |
| **Gestational age at**<br>**birth, weeks** |  | 27.24 ± 1.82 |
| **Birth weight, g** |  | 962.54 ± 263.94 |
| **NICU length of stay,**<br>**days** |  | 88.52 ± 33.99 |
| **Age at BASC-3**<br>**assessment, years** |  | 4.10 ± 0.16 |

Note: Values are presented as N (%) for categorical variables and mean ± standard deviation (SD) for continuous variables. Percentages are based on the total analytic sample of 226 participants. Data were missing for maternal education (n = 4), socioeconomic status (n = 4), birth weight (n = 2), and age at BASC-3 assessment (n = 6). Low socioeconomic status was defined as Hollingshead Index level 5.
Abbreviations: BASC-3, Behavior Assessment System for Children, Third Edition; GED, General Educational Development; NICU, neonatal intensive care unit; SD, standard deviation.

### Association of environmental metabolites and behavioral outcomes

Among the 30 environmental metabolites tested, 2-(Acetylamino)-3-(phenylthio) propanoic acid showed consistent positive associations across all BASC-3 behavioral domains. Higher levels of this metabolite were associated with higher overall behavior problems (β = 1.42, *SE* = 0.52, *p* = 0.006), as well as higher externalizing (β = 1.50, SE = 0.54, p = 0.006), and internalizing behavior problems (β = 1.34, SE = 0.58, p = 0.021) (supplement table 1-3).

Additionally, Diethofencarb was positively associated with overall behavior problems (β = 0.34, SE = 0.14, p = 0.018) and externalizing behaviors (β = 0.39, SE = 0.15, p = 0.010). While protocatechuic acid was inversely associated with internalizing behaviors (β = −0.68, SE = 0.31, p = 0.027). However, these associations were not statistically significant at FDR < 5%.

### Association of environmental metabolites and endogenous metabolites

To investigate the underlying biological processes that may be associated by these behavior-associated environmental metabolites, we performed MWAS for each of these three environmental metabolites and 16,958 endogenous metabolites. Models were adjusted for race, sex, socioeconomic status, study site, gestational age at birth, urinary creatinine, and 10 principal components.

The three environmental metabolites demonstrated widespread associations with endogenous metabolic features. Specifically, 2-(Acetylamino)-3-(phenylthio) propanoic acid was associated with 151 endogenous metabolites, Diethofencarb was associated with 131 endogenous metabolites, and Protocatechuic acid was associated with 530 endogenous metabolites at an FDR threshold of < 0.05 (Figure 1).

**Figure 1.**
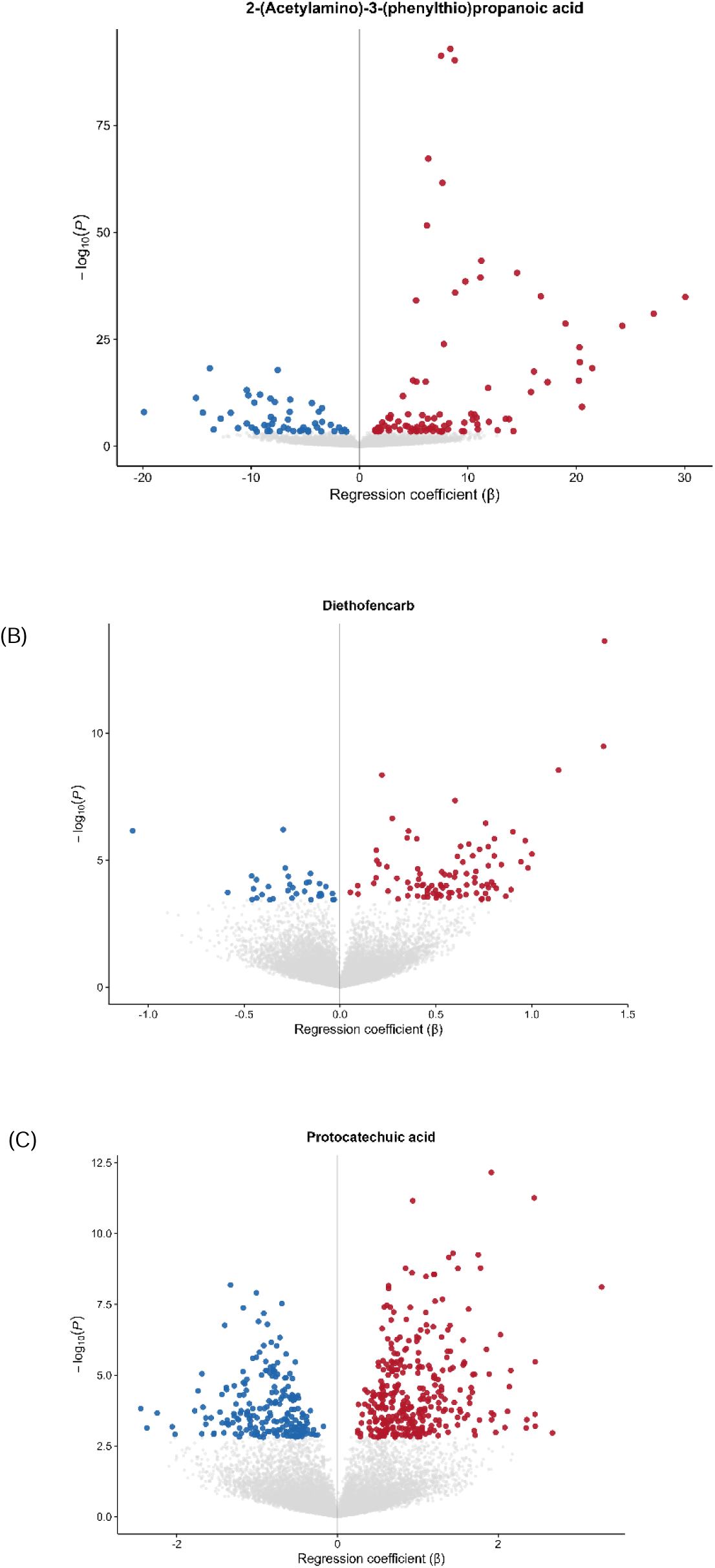
Metabolome-wide associations of behavior-associated environmental metabolites with endogenous metabolic features in children born very preterm. Volcano plots show associations between (A) 2-(Acetylamino)-3-(phenylthio)propanoic acid, (B) Diethofencarb, and (C) Protocatechuic acid and endogenous metabolic features measured in urine at age 4 years. The x-axis represents the regression coefficient (β), and the y-axis represents −log10(P). Features meeting a false discovery rate (FDR) threshold of <0.05 are highlighted according to the direction of association, with positive associations shown in red and negative associations in blue; nonsignificant features are shown in gray

### Pathway Analysis

Pathway enrichment analyses were conducted using endogenous metabolites significantly associated with each of the three behavior-associated environmental metabolites (Figure 2).

**Figure 2.**
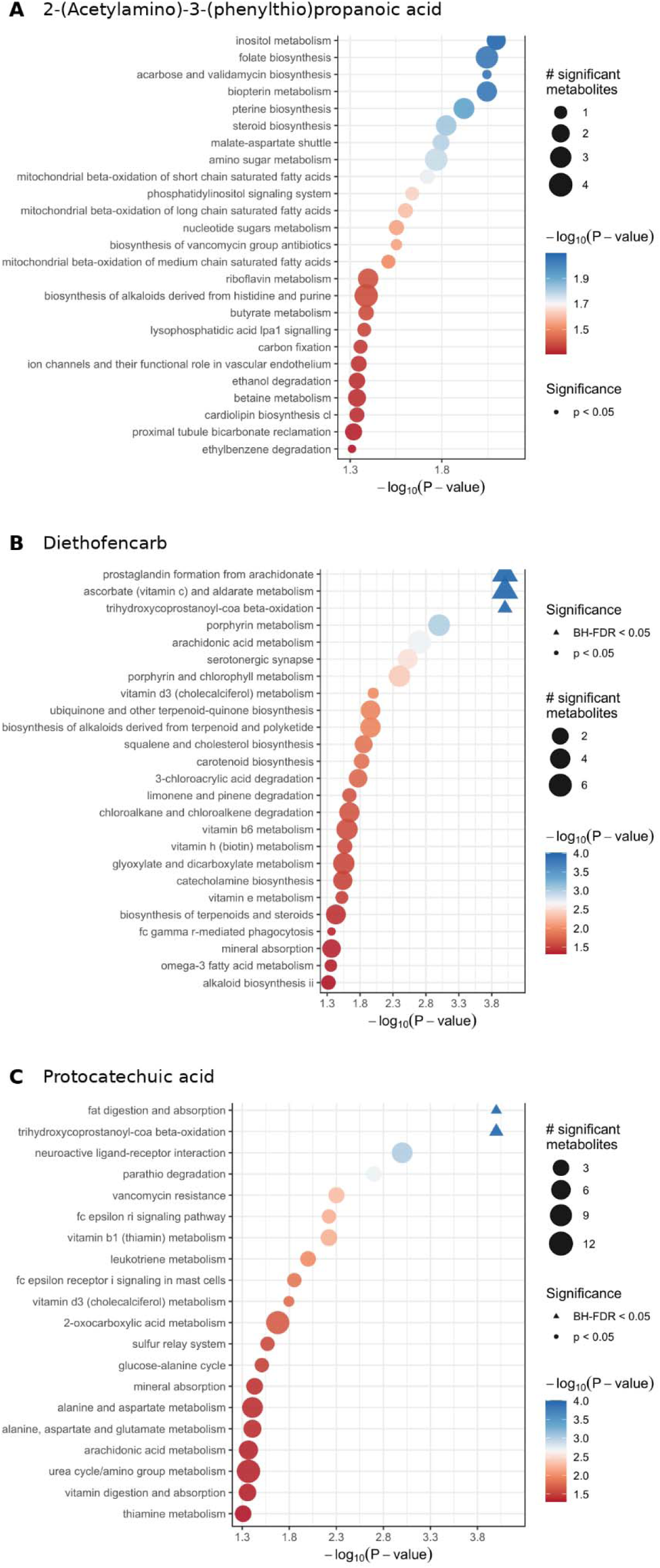
Pathway enrichment associated with behavior-associated environmental metabolites. Pathway enrichment analyses were conducted using endogenous metabolic features significantly associated with (A) 2-(Acetylamino)-3-(phenylthio)propanoic acid, (B) Diethofencarb, and (C) protocatechuic acid. The x-axis represents −log (*P*-value), and point size represents the number of significant metabolites contributing to each pathway. Triangles indicate pathways that remained significant after false discovery rate correction (FDR < 0.05), while circles indicate pathways meeting nominal significance (*P* < 0.05).

For 2-(Acetylamino)-3-(phenylthio)propanoic acid, 25 pathways were significantly enriched at nominal *p* < 0.05 and none of them passed FDR <0.05. Among these, pathways emerged that were primarily related to cellular energy metabolism, lipid metabolism, and micronutrient-dependent processes. Enriched pathways included mitochondrial β-oxidation of short, medium, and long-chain fatty acids, butyrate metabolism, riboflavin metabolism, folate biosynthesis, biopterin metabolism, and inositol metabolism. Additional enrichment of phosphatidylinositol signaling, cardiolipin biosynthesis, amino sugar metabolism, and the malate-aspartate shuttle further highlighted pathways involved in mitochondrial function, cellular signaling, and intermediary metabolism.

For Diethofencarb, 22 pathways were significantly enriched at nominal p < 0.05 and 3 remained significant at FDR<0.05. Among these, pathways primarily related to lipid and fatty acid metabolism, inflammatory signaling, vitamin metabolism, and neurotransmitter-related processes. The FDR-significant pathways included prostaglandin formation from arachidonate, ascorbate and aldarate metabolism, and trihydroxycoprostanoyl-CoA β-oxidation. Broader enrichment patterns among the nominally-significant pathways also included arachidonic acid metabolism, omega-3 fatty acid metabolism, serotonergic synapse, catecholamine biosynthesis, and pathways involving vitamins B6, D3, and E and biotin, highlighting potential involvement of inflammatory lipid signaling, antioxidant processes, and neuroactive metabolism. The three pathways that remained significant at FDR < 0.05 were related to inflammatory lipid signaling (prostaglandin formation from arachidonate), antioxidant metabolism (ascorbate and aldarate metabolism), and fatty acid catabolism (trihydroxycoprostanoyl-CoA β-oxidation).

For Protocatechuic acid, 17 pathways were significantly enriched at p < 0.05 and 2 remained significant at FDR<0.05. Among these, pathways were enriched related to lipid metabolism, amino acid metabolism, neuroactive signaling, and immune-related processes. Fat digestion and absorption and trihydroxycoprostanoyl-CoA β-oxidation were among the pathways that remained significant after multiple-testing correction.

Broader enrichment patterns included alanine, aspartate and glutamate metabolism, the urea cycle/amino group metabolism, the glucose-alanine cycle, arachidonic acid and leukotriene metabolism, and neuroactive ligand-receptor interaction, as well as pathways related to vitamin metabolism. The pathways that remained significant at FDR < 0.05 were related to fat digestion and absorption and trihydroxycoprostanoyl-CoA β-oxidation.

## Discussion

In this study of children born very preterm, we identified three environmentally relevant metabolites associated with behavior problems at age 4 and demonstrated that these metabolites were linked to widespread alterations in endogenous metabolic pathways. Together, our findings provide new insight into potential biological mechanisms through which environmental exposures may influence neurobehavioral outcomes in children born very preterm.

Children born very preterm are particularly susceptible to adverse neurodevelopmental outcomes because of disrupted fetal development, prolonged neonatal intensive care exposure, inflammation and oxidative stress [25–27]. Our findings suggest that environmental exposures during early-childhood are related to differential behavioral vulnerability, potentially through perturbed biological activities. Previous studies have shown that systemic inflammation and oxidative stress are important contributors to impaired neurodevelopment in preterm infants [28], and environmental chemicals can also disrupt these and related biological systems including inflammatory, endocrine, oxidative stress, and mitochondrial pathways [9]. The metabolic pathways that we identified in association with behavioral-associated chemicals, align closely with the biological processes already implicated in neurodevelopment.

2-(Acetylamino)-3-(phenylthio)propanoic acid (phenylmercapturic acid), was positively and consistently associated with overall, externalizing, and internalizing behavior problems. Phenylmercapturic acid is a urinary metabolite of benzene and is commonly used as a biomarker of benzene exposure [29]. Benzene is a widespread environmental contaminant with multiple potential sources, including gasoline and gasoline vapors, motor vehicle exhaust, cigarette smoke, industrial emissions, and other combustion-related sources [30]. Thus, the observed association may reflect a relationship between benzene exposure and behavioral problems. The associated endogenous metabolic pathways were primarily related to cellular energy production, fatty acid metabolism, mitochondrial function, and micronutrient-dependent metabolism. Enrichment of mitochondrial β-oxidation pathways involving short-, medium-, and long-chain fatty acids, together with butyrate metabolism, cardiolipin biosynthesis, and the malate-aspartate shuttle, is related to mitochondrial substrate utilization and energy-generating processes [31–33]. Mitochondria play a central role in brain development because of the high energy requirements of neuronal differentiation, synapse formation, and neurotransmission. Altered mitochondrial energy metabolism has been implicated in neurodevelopmental and behavioral conditions, particularly in populations exposed to early-life physiological stressors [34–36]. Additional enrichment of riboflavin, folate, biopterin, inositol, amino sugar, and phosphatidylinositol signaling pathways further supports the possibility that this metabolite is associated with broader perturbations in cellular metabolism and signaling. Riboflavin and folate are essential cofactors for redox regulation, one-carbon metabolism, and mitochondrial function[37, 38]. Phosphatidylinositol signaling and cardiolipin metabolism are also important for membrane integrity, intracellular signaling, and mitochondrial function [39, 40]. Our observation that 2-(Acetylamino)-3-(phenylthio)propanoic acid was associated with metabolic pathways relevant to mitochondrial function, energy metabolism, and oxidative stress suggests that children born very preterm may be particularly vulnerable to the effects of environmental chemicals that perturb some of these same biological processes.

Diethofencarb, a phenyl-carbamate fungicide [41], was associated with overall and externalizing behavior problems and demonstrated associations with multiple endogenous metabolic pathways, including cytochrome P450 metabolism, steroid hormone biosynthesis, phenylalanine metabolism, tryptophan metabolism, and androgen and estrogen metabolism. Diethofencarb was developed primarily for the control of fungicide-resistant plant pathogens, particularly *Botrytis cinerea* (gray mold), in crops such as fruits, grapes, and vegetables. Although diethofencarb is not registered for agricultural use in the United States, the U.S. Environmental Protection Agency has established an import tolerance for residues on bananas, and diethofencarb residues have also been identified in imported food products through U.S. food surveillance. Thus, dietary exposure through imported produce represents one possible source of diethofencarb in this population; however, because dietary intake and specific sources of exposure were not assessed, the origin of the detected metabolite cannot be determined from the present study. Prior epidemiological studies have linked pesticide exposure with impaired neurobehavioral development, including behavioral problems, attention deficits, and cognitive dysfunction in children [42]. Experimental evidence [43] [44] further suggests that pesticide-related compounds may influence neurodevelopment through oxidative stress, endocrine disruption, mitochondrial dysfunction, and altered neurotransmitter signaling [45–47]. The strongest enrichment was observed for prostaglandin formation from arachidonate, arachidonic acid metabolism, ascorbate and aldarate metabolism, and trihydroxycoprostanoyl-CoA β-oxidation, which remained significant following multiple-testing correction. These pathways point to potential alterations in inflammatory lipid mediator production, antioxidant-related metabolism, and fatty acid oxidation [48, 49]. Arachidonic acid metabolism and prostaglandin synthesis are biologically relevant to neurodevelopment because arachidonic acid and its eicosanoid derivatives contribute to inflammatory and immune signaling, vascular regulation, and neuronal membrane and synaptic processes [50, 51].

Protocatechuic acid demonstrated inverse associations with internalizing behaviors and was associated with the largest number of endogenous metabolic features. Protocatechuic acid is a naturally occurring phenolic compound with known antioxidant and anti-inflammatory properties and is commonly derived from dietary polyphenols found in fruits, vegetables, and plant-based foods [52, 53]. Previous studies have shown that protocatechuic acid can reduce oxidative stress, modulate inflammatory signaling, and exert neuroprotective effects through regulation of antioxidant defense pathways and mitochondrial function [54, 55]. hese properties may be particularly relevant for children born very preterm, who experience elevated oxidative stress and inflammatory burden during early development. Consistent with this potential protective role, our pathway enrichment analysis identified alanine, aspartate and glutamate metabolism, urea cycle/amino group metabolism, and the glucose-alanine cycle, pathways involved in amino acid utilization, cellular energy metabolism, redox regulation, and neurotransmitter homeostasis [56, 57]. Modulation of these pathways may reflect metabolic processes that support antioxidant defenses and help regulate inflammatory and oxidative stress responses. Thus, rather than indicating adverse metabolic disruption, these associations may represent biological pathways through which protocatechuic acid or related dietary exposures contribute to a more favorable metabolic environment relevant to behavioral development. Together, these findings are consistent with, but do not establish, a potential protective role for protocatechuic acid-associated metabolic processes in children born very preterm.

The three behavior-associated environmental metabolites tended to converge on several interconnected biological domains: mitochondrial energetics, oxidative stress and neurotransmission. This convergence may be particularly relevant for children born very preterm, who experience substantial oxidative and inflammatory stress during a period of rapid brain development. Additionally, mitochondrial function and redox homeostasis play important roles in brain maturation throughout childhood [13].

Environmental exposures that perturb these same interconnected biological systems may therefore be especially important in shaping neurodevelopmental trajectories following very preterm birth. Although these findings do not establish causal mechanisms, they identify biologically plausible pathways that warrant further investigation as potential links between environmental exposures and neurobehavioral outcomes in children born very preterm.

Our study has several strengths. First, we examined environmental exposure metabolites and endogenous metabolites simultaneously within the same high-resolution metabolomic platform, allowing for integrated analysis of external exposures and internal biological responses. Second, the use of untargeted metabolomics enabled comprehensive assessment of a broad range of metabolites without restricting analyses to a limited number of predefined exposure classes. Third, the NOVI cohort provides a unique opportunity to study children born very preterm, a population with unique biological vulnerabilities and elevated risk for neurodevelopmental impairments. Several limitations should also be considered. Urinary metabolites and behavioral outcomes were both measured at a single time point at age 4, which may not fully capture long-term or cumulative exposure patterns or temporal associations. In addition, urine samples were collected under non-fasting conditions, and dietary intake or recent nutritional exposures may have influenced measured metabolite levels. While our focus on children born very preterm is a strength, it also limits generalizability of our findings to their term-born counterparts. Last, our sample size (N=226) was moderate, and individual environmental metabolites that were associated with behavior did not yield FDR q-values < 0.05 and thus these findings should be viewed as hypothesis generating.

## Conclusion

In this MWAS of children born very preterm, we identified associations between environmentally relevant metabolites and neurobehavioral outcomes and found that these behavior-associated metabolites were linked to broad perturbations in endogenous metabolic pathways. Although the associations with behavioral outcomes did not remain significant after multiple testing correction, downstream metabolic analyses identified signals involving mitochondrial energy metabolism, fatty acid oxidation, inflammatory lipid mediators, amino acid metabolism, micronutrient-dependent pathways, xenobiotic processing, and neuroactive signaling. These findings suggest that environmental exposures may influence behavioral development through interconnected metabolic processes that may be particularly relevant in children born very preterm. Larger longitudinal studies incorporating repeated exposure assessment, targeted metabolite confirmation, dietary characterization, and additional molecular data are needed to validate these findings and clarify the biological pathways linking environmental exposures with neurobehavioral development.

## Data Availability

All data produced in the present study are available upon reasonable request to the authors

## REFERENCES

1. Rauh, V.A. and A.E. Margolis, Research Review: Environmental exposures, neurodevelopment, and child mental health - new paradigms for the study of brain and behavioral effects. J Child Psychol Psychiatry, 2016. 57(7): p. 775–93.

2. James, A.A. and O.S. KL, Environmental chemical exposures and mental health outcomes in children: a narrative review of recent literature. Front Toxicol, 2023. 5: p. 1290119.

3. Ferguson, K.K., M.S. O’Neill, and J.D. Meeker, Environmental contaminant exposures and preterm birth: a comprehensive review. J Toxicol Environ Health B Crit Rev, 2013. 16(2): p. 69–113.

4. Santos, J., S.E. Pearce, and A. Stroustrup, Impact of hospital-based environmental exposures on neurodevelopmental outcomes of preterm infants. Curr Opin Pediatr, 2015. 27(2): p. 254–60.

5. Braun, J.M., Early-life exposure to EDCs: role in childhood obesity and neurodevelopment. Nat Rev Endocrinol, 2017. 13(3): p. 161–173.

6. Dewey, D. and M.H. Soomro, Prenatal and childhood exposures to heavy metals and their associations with child cognition, motor skills, behaviour and mental health. Essays Biochem, 2025. 69(3): p. 199–240.

7. Viglietti, P.G., et al., Postnatal exposure to current use pesticides and neurodevelopmental outcomes in children and adolescents: A systematic review. Neurotoxicology, 2026. 116: p. 103542.

8. Castagna, A., et al., Air pollution and neurodevelopmental skills in preschool- and school-aged children: A systematic review. Neurosci Biobehav Rev, 2022. 136: p. 104623.

9. Bennett, D.H., et al., Environmental exposures to pesticides, phthalates, phenols and trace elements are associated with neurodevelopment in the CHARGE study. Environ Int, 2022. 161: p. 107075.

10. Oh, J., et al., Early childhood exposure to environmental phenols and parabens, phthalates, organophosphate pesticides, and trace elements in association with attention deficit hyperactivity disorder (ADHD) symptoms in the CHARGE study. Res Sq, 2023.

11. Rock, K.D. and H.B. Patisaul, Environmental Mechanisms of Neurodevelopmental Toxicity. Curr Environ Health Rep, 2018. 5(1): p. 145–157.

12. Nadal, A., et al., Endocrine-disrupting chemicals and the regulation of energy balance. Nat Rev Endocrinol, 2017. 13(9): p. 536–546.

13. Sirois, C.L., et al., Mitochondrial dynamics in neurodevelopment and neurodevelopmental disorders. Nat Rev Neurosci, 2026. 27(5): p. 307–326.

14. Li, Z., et al., Unraveling the Molecular Links between Fine Particulate Matter Exposure and Early Birth Risks in African American Mothers: A Metabolomics Study in the Atlanta African American Maternal-Child Cohort. Environ Sci Technol, 2025. 59(22): p. 10905–10918.

15. Hoffman, S.S., et al., Impact of prenatal phthalate exposure on newborn metabolome and infant neurodevelopment. Nat Commun, 2025. 16(1): p. 2539.

16. Gabrielli, J., Y. Jackson, and S. Brown, Measurement of Behavioral and Emotional Outcomes of Youth in Foster Care: Investigation of the Roles of Age and Placement Type. J Psychopathol Behav Assess, 2015. 37(3): p. 422–431.

17. Bradstreet, L.E., et al., Using the BASC-2 Parent Rating Scales to Screen for Autism Spectrum Disorder in Toddlers and Preschool-Aged Children. J Abnorm Child Psychol, 2017. 45(2): p. 359–370.

18. Reynolds C, K.R., Behavior Assessment System for Children, Third Edition. 2015, Circle Pines, MN.

19. Sato, J., et al., Social-Cognitive Network Connectivity in Preterm Children and Relations With Early Nutrition and Developmental Outcomes. Front Syst Neurosci, 2022. 16: p. 812111.

20. Taibl, K.R., et al., Newborn metabolomic signatures of maternal per- and polyfluoroalkyl substance exposure and reduced length of gestation. Nat Commun, 2023. 14(1): p. 3120.

21. Li, Y.Y., et al., Untargeted Metabolomics: Biochemical Perturbations in Golestan Cohort Study Opium Users Inform Intervention Strategies. Front Nutr, 2020. 7: p. 584585.

22. Schymanski, E.L., et al., Identifying small molecules via high resolution mass spectrometry: communicating confidence. Environ Sci Technol, 2014. 48(4): p. 2097–8.

23. Broadhurst, D., et al., Guidelines and considerations for the use of system suitability and quality control samples in mass spectrometry assays applied in untargeted clinical metabolomic studies. Metabolomics, 2018. 14(6): p. 72.

24. Tian, L., et al., Metapone: a Bioconductor package for joint pathway testing for untargeted metabolomics data. Bioinformatics, 2022. 38(14): p. 3662–3664.

25. Humberg, A., et al., Preterm birth and sustained inflammation: consequences for the neonate. Semin Immunopathol, 2020. 42(4): p. 451–468.

26. Lembo, C., G. Buonocore, and S. Perrone, Oxidative Stress in Preterm Newborns. Antioxidants (Basel), 2021. 10(11).

27. Hee Chung, E., J. Chou, and K.A. Brown, Neurodevelopmental outcomes of preterm infants: a recent literature review. Transl Pediatr, 2020. 9(Suppl 1): p. S3–S8.

28. Martini, S., et al., Antenatal and Postnatal Sequelae of Oxidative Stress in Preterm Infants: A Narrative Review Targeting Pathophysiological Mechanisms. Antioxidants (Basel), 2023. 12(2).

29. Farmer, P.B., et al., The use of S-phenylmercapturic acid as a biomarker in molecular epidemiology studies of benzene. Chem Biol Interact, 2005. 153-154: p. 97–102.

30. Weisel, C.P., Benzene exposure: an overview of monitoring methods and their findings. Chem Biol Interact, 2010. 184(1-2): p. 58–66.

31. Xavier, J.M., C.M. Rodrigues, and S. Sola, Mitochondria: Major Regulators of Neural Development. Neuroscientist, 2016. 22(4): p. 346–58.

32. Szrok-Jurga, S., et al., The Role of Acyl-CoA beta-Oxidation in Brain Metabolism and Neurodegenerative Diseases. Int J Mol Sci, 2023. 24(18).

33. Zhang, L., et al., Butyrate in Energy Metabolism: There Is Still More to Learn. Trends Endocrinol Metab, 2021. 32(3): p. 159–169.

34. Cheng, A., Y. Hou, and M.P. Mattson, Mitochondria and neuroplasticity. ASN Neuro, 2010. 2(5): p. e00045.

35. Fame, R.M. and M.K. Lehtinen, Mitochondria in Early Forebrain Development: From Neurulation to Mid-Corticogenesis. Front Cell Dev Biol, 2021. 9: p. 780207.

36. Zhao, T., et al., Pain/Stress, Mitochondrial Dysfunction, and Neurodevelopment in Preterm Infants. Dev Neurosci, 2024. 46(5): p. 341–352.

37. Henriques, B.J., et al., Emerging roles for riboflavin in functional rescue of mitochondrial beta-oxidation flavoenzymes. Curr Med Chem, 2010. 17(32): p. 3842–54.

38. Ducker, G.S. and J.D. Rabinowitz, One-Carbon Metabolism in Health and Disease. Cell Metab, 2017. 25(1): p. 27–42.

39. Hammond, G.R.V. and J.E. Burke, Novel roles of phosphoinositides in signaling, lipid transport, and disease. Curr Opin Cell Biol, 2020. 63: p. 57–67.

40. Paradies, G., et al., Cardiolipin and mitochondrial function in health and disease. Antioxid Redox Signal, 2014. 20(12): p. 1925–53.

41. Fujisawa, T., et al., Metabolism of fungicide diethofencarb in grape (Vitis vinifera L.): definitive identification of thiolactic acid conjugated metabolites. J Agric Food Chem, 2003. 51(18): p. 5329–36.

42. Jurewicz, J. and W. Hanke, Prenatal and childhood exposure to pesticides and neurobehavioral development: review of epidemiological studies. Int J Occup Med Environ Health, 2008. 21(2): p. 121–32.

43. Hood, R.B., et al., Pesticide residue intake from fruits and vegetables and alterations in the serum metabolome of women undergoing infertility treatment. Environ Int, 2022. 160: p. 107061.

44. Liang, D., et al., Metabolome-wide association study of the relationship between chlorpyrifos exposure and first trimester serum metabolite levels in pregnant Thai farmworkers. Environ Res, 2022. 215(Pt 2): p. 114319.

45. Kori, R.K., et al., Neurochemical and Behavioral Dysfunctions in Pesticide Exposed Farm Workers: A Clinical Outcome. Indian J Clin Biochem, 2018. 33(4): p. 372–381.

46. Chilipweli, P.M., et al., The role of cytochrome P450 genetic variants in pesticide metabolism and the risk of child neurodevelopment: a systematic review. Front Pediatr, 2025. 13: p. 1651916.

47. Botnaru, A.A., et al., Neurotoxic Effects of Pesticides: Implications for Neurodegenerative and Neurobehavioral Disorders. J Xenobiot, 2025. 15(3).

48. Funk, C.D., Prostaglandins and leukotrienes: advances in eicosanoid biology. Science, 2001. 294(5548): p. 1871–5.

49. Zhitkovich, A., Ascorbate: antioxidant and biochemical activities and their importance for in vitro models. Arch Toxicol, 2021. 95(12): p. 3623–3631.

50. Basak, S., et al., Maternal Supply of Both Arachidonic and Docosahexaenoic Acids Is Required for Optimal Neurodevelopment. Nutrients, 2021. 13(6).

51. Bosetti, F., Arachidonic acid metabolism in brain physiology and pathology: lessons from genetically altered mouse models. J Neurochem, 2007. 102(3): p. 577–86.

52. Zhang, S., et al., Antioxidant Effects of Protocatechuic Acid and Protocatechuic Aldehyde: Old Wine in a New Bottle. Evid Based Complement Alternat Med, 2021. 2021: p. 6139308.

53. Kakkar, S. and S. Bais, A review on protocatechuic Acid and its pharmacological potential. ISRN Pharmacol, 2014. 2014: p. 952943.

54. Vari, R., et al., Protocatechuic Acid Prevents oxLDL-Induced Apoptosis by Activating JNK/Nrf2 Survival Signals in Macrophages. Oxid Med Cell Longev, 2015. 2015: p. 351827.

55. Kelidari, M., et al., The protective effects of protocatechuic acid against natural and chemical toxicants: cellular and molecular mechanisms. Naunyn Schmiedebergs Arch Pharmacol, 2024. 397(8): p. 5597–5616.

56. Chandel, N.S., Amino Acid Metabolism. Cold Spring Harb Perspect Biol, 2021. 13(4).

57. Felig, P., The glucose-alanine cycle. Metabolism, 1973. 22(2): p. 179–207.

